# Reproducibility of 7T MRI Measurements of The Susceptibility and Volume of Hippocampal Subfields

**DOI:** 10.64898/2026.06.15.26355711

**Authors:** O.F Adeyemi, Olivier Mougin, Catarina Rua, Christopher T. Rodgers, Penny Gowland, Akram A. Hosseini, Richard Bowtell

**Affiliations:** Sir Peter Mansfield Imaging Centre, School of Physics and Astronomy, University of Nottingham, Nottingham, NG7 2RD; Medical Physics, RRPPS, University Hospital Birmingham NHS Foundation Trust, 63 Melchett Rd, Birmingham B30 3HP; Department of Physics, University of Abuja, Gwagwalada FCT, Nigeria; Wolfson Brain Imaging Centre, University of Cambridge, Cambridge Biomedical Campus, Cambridge, CB2 0QQ; Antaros Medical, 431 53 Mölndal, Sweden; Department of Neurology, Nottingham University Hospitals NHS Trust, Queen’s Medical Centre, Derby Road, Nottingham NG7 2UH

**Keywords:** QSM, susceptibility, hippocampus, hippocampal subfields, reproducibility

## Abstract

**PURPOSE:** The UK7T travelling head dataset was used to characterise the reproducibility of 7T measurements of the susceptibility of the hippocampal subfields, focusing on the Cornu Ammonis (CA1, CA2 and CA3), dentate gyrus (DG), subiculum (SUB), tail of the hippocampus (TAIL) and entorhinal cortex (ERC).

**METHODS:** Susceptibility maps were created from whole-brain 3D single-echo GRE data (TE=20 ms; 0.7 mm isotropic resolution) using Multi-Scale Dipole Inversion. Automatic Segmentation of Hippocampal Subfields (ASHS) was applied to high resolution T1– and T2-weighted images for segmentation. The mean magnetic susceptibility and volume of hippocampal subfields was evaluated in 50 data sets, comprising 5 repeat acquisitions on 10 healthy participants (age 32±6 years; 3 female).

**RESULTS:** Averaging over subjects, susceptibility values spanned an 18ppb range over the hippocampus (ranging from –13.3ppb in DG to 4.7ppb in ERC). Susceptibility values in the larger hippocampal subfields showed a consistent pattern of variation across subjects, being generally more positive in ERC and SUB than in CA1 and more positive in CA1 than in DG and TAIL. The standard deviation of subfield susceptibilities over subjects ranged from 8.2ppb in the TAIL to 1.7ppb in CA1, and the average standard deviation across repeated measurements, which ranges from 1.7 to 4 ppb, was less than half of the inter-participant standard deviation in all subfields. Susceptibility values in the smaller subfields (CA2 and CA3) were more variable, but ICC(2,k) values for all subfields were >0.82.

**CONCLUSION:** The reported data characterises the variation and reproducibility of hippocampal subfield susceptibility measurements at 7T.

## INTRODUCTION

MRI-based quantitative susceptibility mapping (QSM) provides measurements of the spatial variation of the magnetic susceptibility within tissue (1). QSM measurements are particularly sensitive to the concentration of iron (2) and the appearance of susceptibility maps of the brain is largely dominated by the spatial variation of non-haeme iron content (3). Higher field strength increases the sensitivity of gradient echo images to susceptibility variation, which along with the increase in intrinsic signal-to-noise ratio, means that QSM data can be acquired at higher spatial resolution. QSM at 7T consequently provides a powerful approach for characterizing brain iron concentration at sub-millimetre resolution (4,5).

Tissue iron is vital for oxygen transport, neurotransmitter synthesis, and myelin formation, but excess iron can generate reactive oxygen species that drive oxidative stress and damage neurons (6). Longstanding post-mortem and in vivo measurements have revealed abnormal changes in regional brain iron content across a range of neurodegenerative diseases, including Alzheimer’s (7,8) and Parkinson’s disease (9–11), suggesting an association between iron content and the pathology of neurodegeneration. In recent years QSM has been increasingly applied to the investigation of neurodegenerative disease (12), and in the case of Alzheimer’s disease (13), several studies have shown increased magnetic susceptibility in cortical and sub-cortical regions, with some correlation with level of cognitive impairment (14–16).

Located within the medial temporal lobes, the hippocampus plays a central role in multiple cognitive functions including memory and spatial reasoning (17). It is formed from subfields, that can be identified based on their cellular organization, with the anatomical differences related to subfield-specific functional contributions. The hippocampus, and in particular its CA1 subfield (CA = cornu ammonis), is vulnerable to metabolic stress, a feature evident in several acute and subacute neurological conditions such as cerebral ischaemia, limbic encephalitis, hypoglycaemic encephalopathy, and transient global amnesia (18). AD is characterized by early and prominent degeneration of the hippocampus (19) and hippocampal volumetry has therefore become a valuable imaging marker for disease monitoring, follow-up and treatment adjustment (20–22). Several QSM-based studies have shown increased susceptibility in the whole hippocampus in AD and mild cognitive impairment (MCI) (23,24), consistent with elevated iron content, but this increase was not reported in all studies (12) considered in a recent systematic review. This discrepancy may be because of the small size of the hippocampus and the relatively coarse resolution used in earlier studies, which precluded analysis of regional variation in hippocampal susceptibility. The higher spatial resolution available at 7T overcomes this issue and high resolution QSM at 7T has recently been used to evaluate changes in the susceptibility of different hippocampal subfields in participants with MCI and early AD compared to healthy controls (25–27). Reliable identification of parts per billion (ppb) differences in magnetic susceptibility in the mm-scale subfields of the hippocampus, requires the implementation of QSM with a high degree of reproducibility. Previous studies of QSM reproducibility have generally focused on measurements in regions of interest of larger size than the hippocampal subfields (5,28,29). In this study, which was carried out in preparation for measurements of longitudinal changes in the hippocampal subfields in AD, we used the UK7T travelling head study dataset (30) which includes high-resolution T_2_*-weighted gradient echo for QSM, along with T_1_-weighted structural MPRAGE and T_2_-weighted TSE data,, to characterise the reproducibility of 7T susceptibility measurements in the subfields of the hippocampus, specifically focusing upon the Cornu Ammonis 1 (CA1, CA2 and CA3), the dentate gyrus (DG), the subiculum (SUB), the tail of the hippocampus (TAIL) and the entorhinal cortex (ERC). The reproducibility of measurements of the volumes of the hippocampal subfields in this dataset was also characterised.

## METHODS

### IMAGING PROTOCOL

Ten healthy volunteers (age 32 ± 6 years; 3 females, 7 male) were scanned in 2018 with informed consent and in compliance with ethical approval (HBREC.2017.08). Two subjects were recruited at each site, scanned five times at their “home” site and once at each of the other four sites, as previously described (30). In the analysis reported here we focus on the 5 repeated scans acquired at the “home” sites. Details of the scanners in use at the time of the measurements are repeated here in Table S1 in the Supporting Information. For each subject, all scans were acquired in separate sessions and the median time over which scans were acquired was 59 days (range 3–71 days) (5).

QSM was based on the acquisition of whole brain 3D single-echo GRE data with TE=20 ms, TR=43 ms; FOV = 224 × 224 × 224 mm^3^, voxel size =0.7 ×0.7 ×0.7 mm^3^, bandwidth per pixel = 70Hz and acquisition time =12:38 minutes (30). High resolution T1– and T2-weighted images were also acquired in each session to allow segmentation of the hippocampus. These comprised a whole-brain T1-weighted 3D-MP2RAGE image data set (inversion times = 725/2150 ms; TE=2.64 ms; TR= 3500 ms, FOV = 224 × 224 × 224 mm^3^, voxel size =0.8 × 0.8 ×0.8 mm^3^ and acquisition time =7:51 minutes) (30) and a T2-weighted TSE image data set spanning the hippocampus (TE=76 ms; TR= 8020 ms, FOV = 224 × 224 × 55 mm^3^, voxel size =0.4 × 0.4 ×1.0 mm^3^ and acquisition time =4:32 minutes). The slice orientation for the TSE images was chosen so that the slices ran orthogonal to the long axis of the body of the hippocampus.

### QSM RECONSTRUCTION

Susceptibility maps of the brain were created from the GRE images using Multi-Scale Dipole Inversion (MSDI) available as QSMbox v2.0. (31). Background field removal was optimized through a dual-stage approach, initially applying a Laplacian boundary value approach (32) followed by a variable spherical mean value method (33), to mitigate potential artifacts from imperfect field removal. The subsequent field-to-susceptibility inversion utilized a regularization factor of λ = 10^2.7^ which was previously validated for high-resolution 7T imaging via L-curve analysis (31).

### AUTOMATIC SEGMENTATION OF HIPPOCAMPAL SUBFIELDS (ASHS)

Hippocampal segmentation was based on application of the open-source, Automatic Segmentation of Hippocampal Subfields (ASHS Version 2.0) software (34) to the T1– and T2-weighted images acquired in each of the five scan sessions, using a pipeline that combines multi-atlas label fusion and learning-based error correction (34) (35). Hippocampal subfields delineated using this approach based on the UMC Utrecht 7T ASHS Atlas were the Cornu Ammonis (CA) subfields, CA1, CA2 and CA3, plus the DG, SUB, ERC, and TAIL. The mean magnetic susceptibility (ppm) and volume (mm^3^) of each hippocampal subfield were evaluated for each scan. Susceptibility values were effectively measured relative to the average whole-brain susceptibility.

The co-registration of structural and quantitative data followed a two-stage pipeline. First, whole-brain T1-weighted MP2RAGE volumes were aligned to the high-resolution T2-weighted TSE hippocampal slabs using the automated multi-stage registration framework within ASHS. Second, to extract susceptibility values, the ASHS-derived subfield masks were mapped from the TSE space into the native GRE (QSM) space using FSL (36). This was achieved by calculating a rigid-body transformation between the TSE and the GRE magnitude images using FSL FLIRT (37). All hippocampal subfield masks were resampled into the QSM image space using nearest-neighbour interpolation to preserve label integrity, and mean susceptibility values and volumes were subsequently extracted using the fslstats utility (36). We did not find any significant differences between the left and right hippocampus in the volume or susceptibility of subfields and consequently report the volume and susceptibility of the combination of left and right sub-fields here.

## RESULTS

50 datasets were considered in this analysis, corresponding to the five repeated measurements acquired for each of the 10 subjects in the UK7T Travelling Heads study. Typical results of the segmentation are shown in Figure 1, which displays five repeat measurements made on one subject (Subject 7) overlaid on the corresponding TSE data. High quality susceptibility maps were obtained by applying the MSDI approach to the 0.7 mm isotropic resolution GRE data (5). Figure 2 shows single coronal slices through the hippocampus in QSM data derived from one of the repeated measurements on each of the 10 subjects, with and without the results of the segmentation overlaid. Figure S1 shows similar axial, coronal and sagittal slices from QSM data acquired in five repeated measurements from one subject (Subject 7).

**Figure 1.**
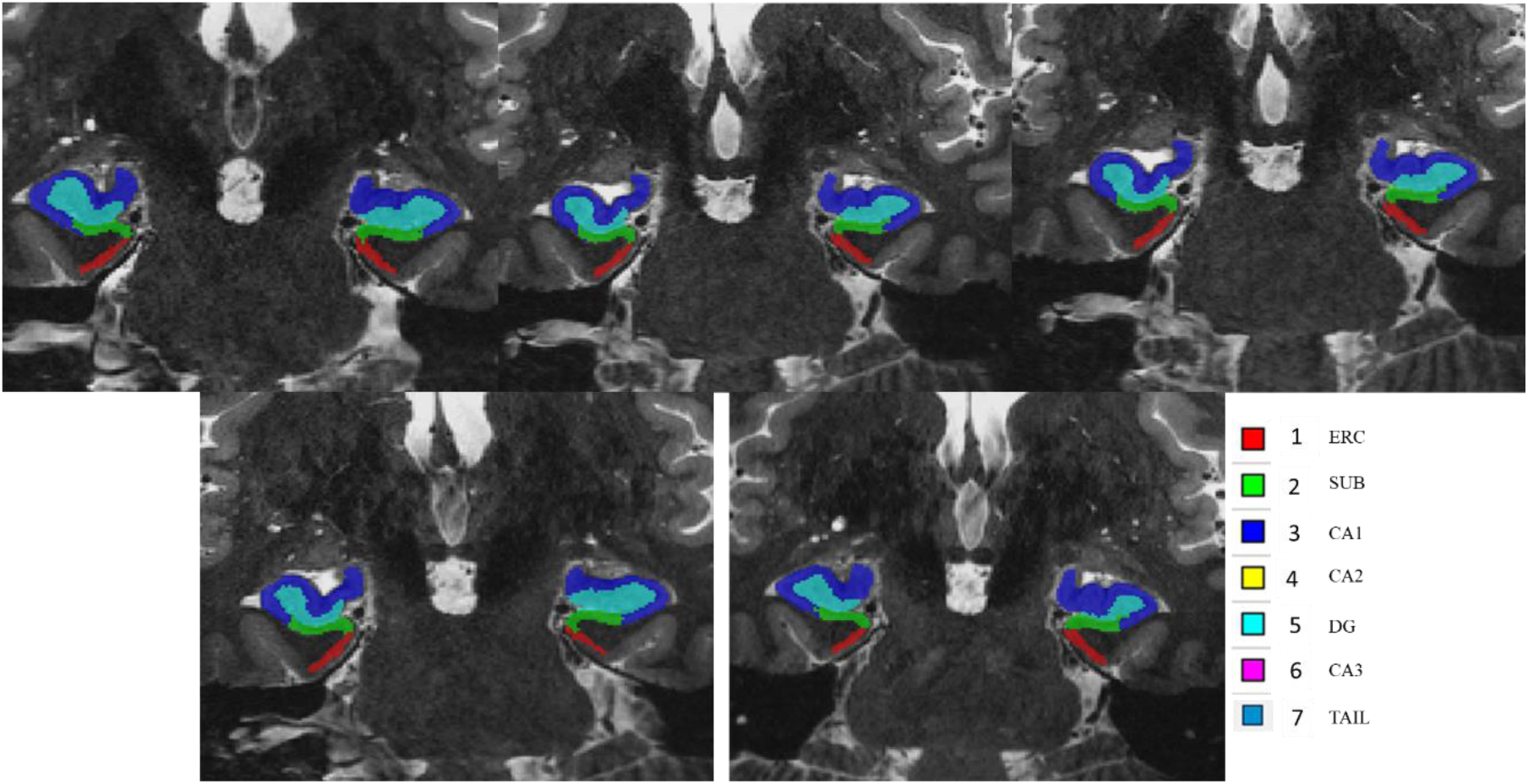
Hippocampal subfield segmentation overlaid on similar slice of coronal TSE images from one participant across the five repeated measurements.

**Figure 2.**
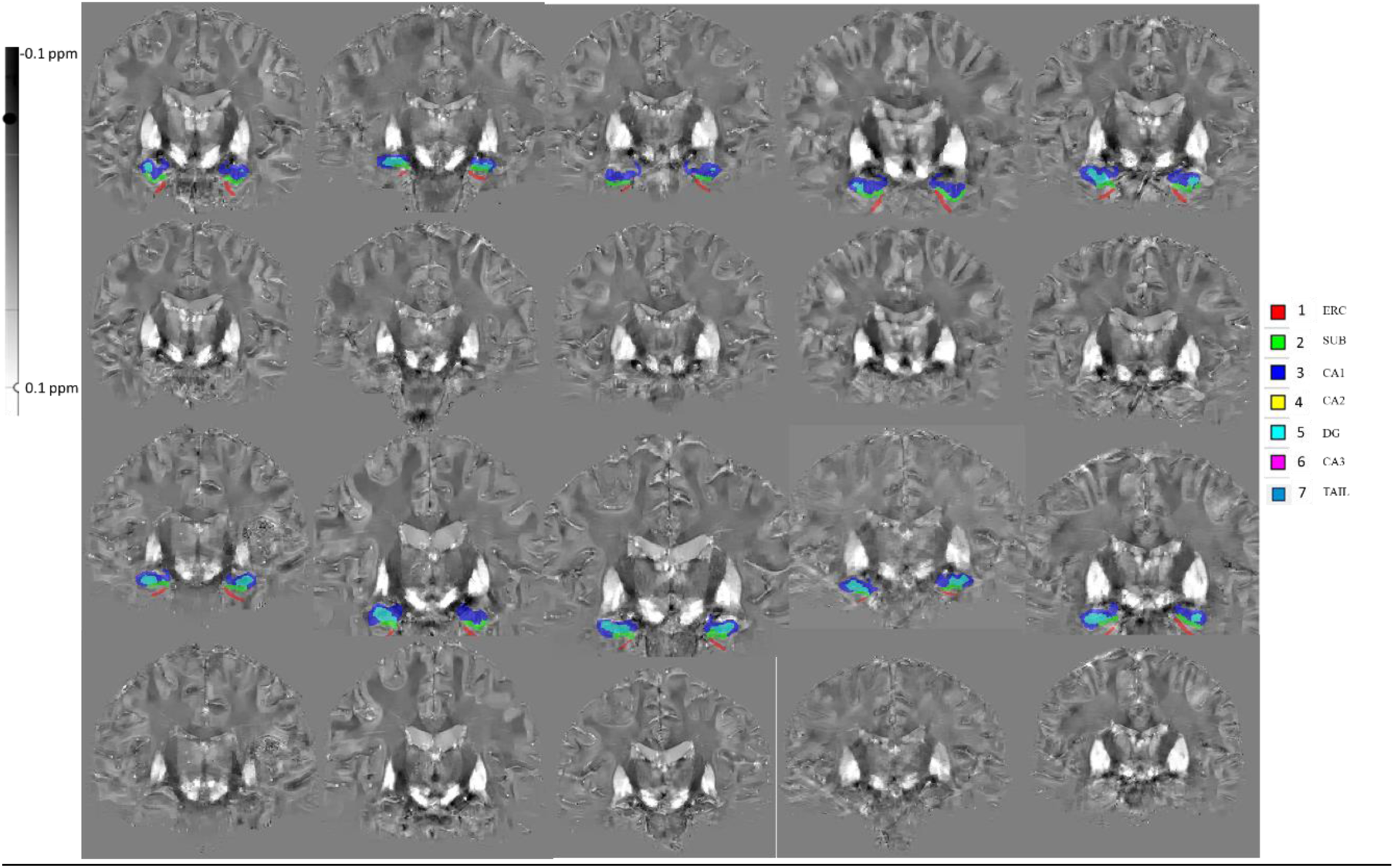
Similar coronal slices drawn from one of the five susceptibility maps acquired from each of the ten participants involved in this study, with and without the ASHS hippocampal segmentations overlaid. Top row: Subjects 1-5 with ASHS overlay; 2^nd^ row: Subjects 1-5 without ASHS overlay; 3^rd^ row: Subjects 6-10 with ASHS overlay; 4^th^ row: Subjects 6-10 without ASHS overlay.

Figure 3 shows plots of the variation of mean susceptibility and volume across the ERC, SUB, CA1, DG and TAIL hippocampal subfields for the 10 participants, along with the standard deviation of the five repeated measurements. Similar plots are shown in Figure 4 for the smaller-volume CA2 and CA3 subfields. Table 2 reports the mean susceptibility and volume measurements across the 10 participants for each hippocampal subfield. We also report the mean, across the 10 participants of the within-subject standard deviations across repeat scans, together with the standard deviations of the mean measurements across participants.

**Figure 3.**
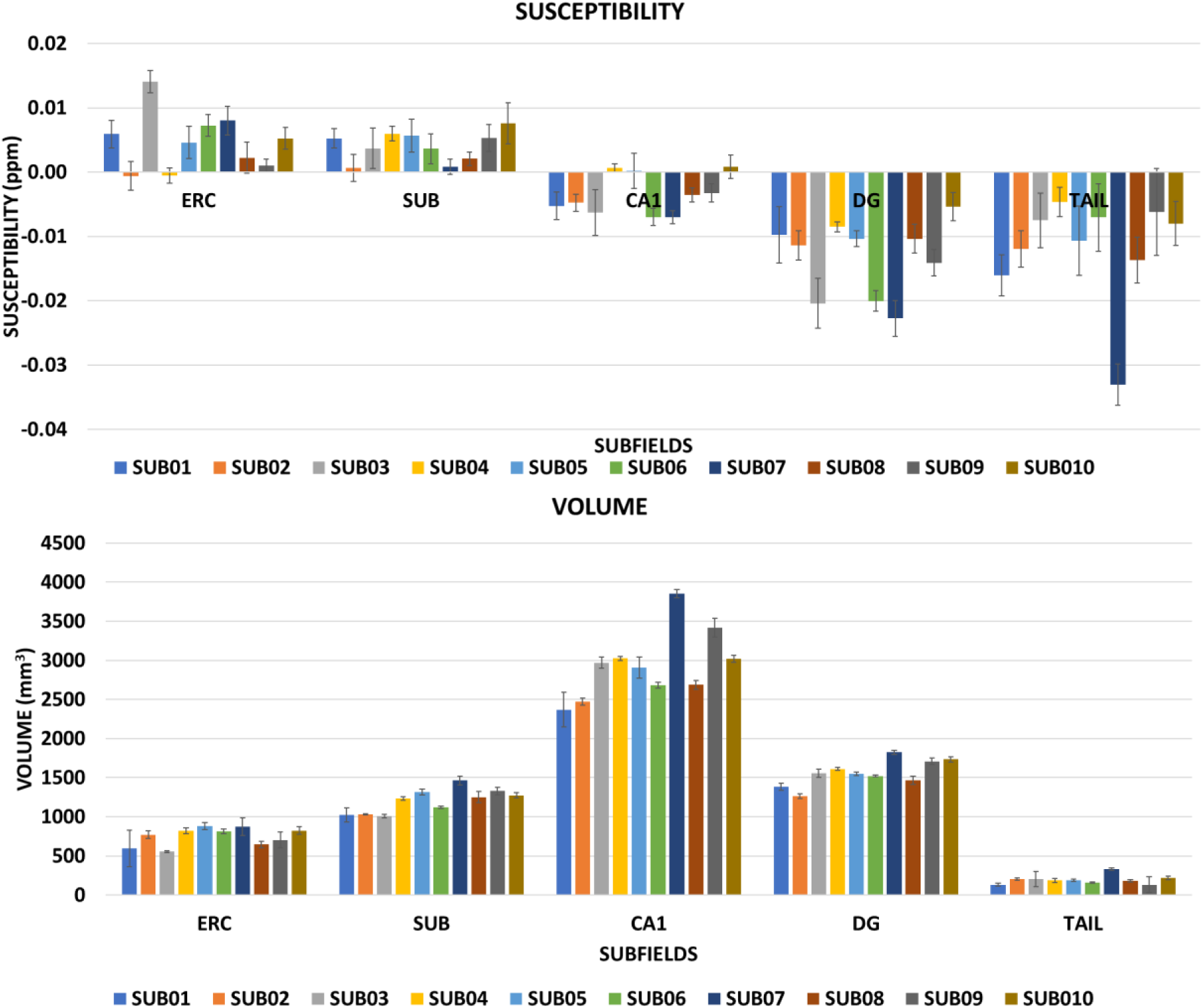
Variation of susceptibility (ppm) and volume (mm^3^) in the larger hippocampal subfields (ERC, SUB, CA1, DG and TAIL) across the 10 participants. Error bars show the standard deviation across repeated measurements.

**Figure 4.**
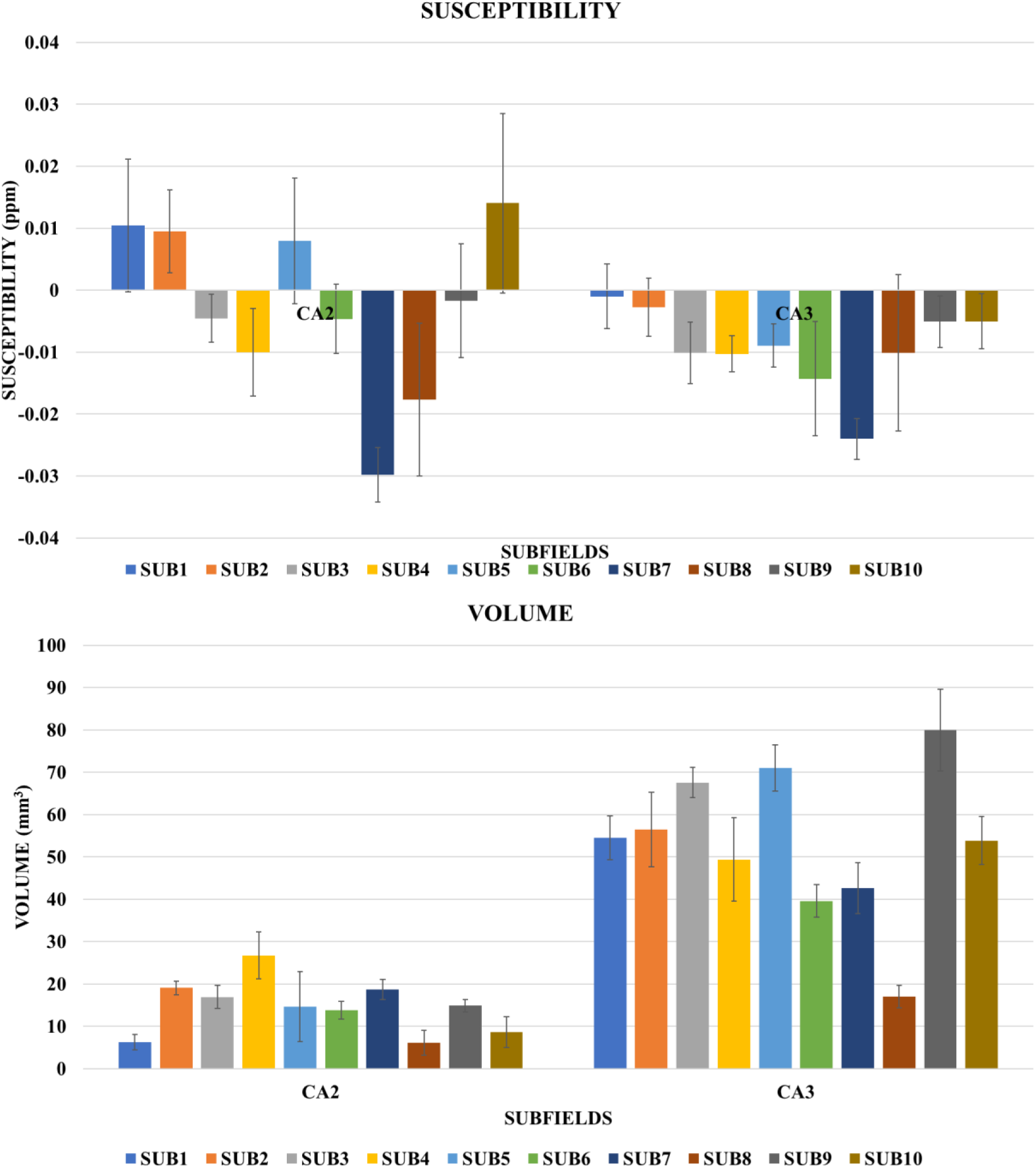
Variation of susceptibility (ppm) and volume (mm^3^) in the smaller hippocampal subfields (CA2 and CA3) across the 10 participants. Error bars show the standard deviation across repeated measurements.

Test–retest reliability across five repeated measurements was also assessed using intraclass correlation coefficients (ICCs). Both single-measure ICC(2,1) and average-measure ICC(2,k) values were calculated and are reported in Table S3 in Supporting Information.

## DISCUSSION AND CONCLUSIONS

The susceptibility values in the larger volume hippocampal subfields show a consistent pattern of variation across subjects, being more positive in the ERC and SUB subfields than in CA1 in all but one subject (Subject 4, for whom CA1 was more positive than ERC), and more positive in CA1 (and in ERC and SUB) than in the DG and TAIL subfields in all subjects (Figure 3). Averaging over subjects, the susceptibility values span 18 ppb, ranging from –13.3 ppb in DG to 4.7 ppb in ERC (Table 1), noting here that referencing is with respect to the average susceptibility within the whole brain mask. The magnitude of the standard deviation across subjects of these subfield susceptibilities ranges from 8.2 ppb in the TAIL to 1.7 ppb in CA1, and in all cases the average standard deviation across repeated measurements is less than half of the inter-participant standard deviation. Figure 4 shows that the CA2 and CA3 subfields which have small average volumes of 15 and 53 mm^3^, respectively, show larger variation in susceptibility across subjects, but the values in CA3 are negative in all subjects (and more negative than the values in CA1 in 8 out of 10 subjects). The greater variability of the measured susceptibility values in the CA2 and CA3 subfields is reflected in the relatively large values of the inter-participant standard deviation (15.6 and 8.5 ppb) and in the standard deviations across repeats (8.4 and 5.5 ppb) reported in Table 1. This likely results from greater sensitivity to segmentation and measurement errors in these small sub-regions (the 15 mm^3^ CA2 subfield volume corresponds to just 44 voxels in the susceptibility map). The ICC(2,1) values for susceptibility measurements (Table S3) ranged from 0.48 (SUB and CA3) to 0.84 (ERC), while ICC(2,k) values were all above 0.82, indicating that the inter-participant variance dominates the variance due to errors across repeated measurements and suggesting that high-resolution QSM at 7T can be used to characterise the variation of subfield susceptibility values between individuals – in this case 10 relatively young subjects with a mean age of 32 years (3 female). The statistical significance of the differences in susceptibility between the different subfields was tested using a one-way repeated ANOVA (setting the alpha value to 0.05), followed by Tukey’s Honest Significant Difference (HSD) test. The results shown in Table S4, indicate that amongst the larger subfields, differences were significant except between ERC and SUB and between DG and TAIL. In the case of the smaller CA2 and CA3 subfields, CA2 did not reach a level of significant difference when compared with the ERC, SUB, or CA1 subfields. Similarly, CA3 did not achieve significance in its difference from the TAIL subfield. These findings may be attributed to the relatively small volume of these specific subfields, which limits the statistical power to detect subtle variations in susceptibility.

**Table 1:** Mean susceptibility and volume across the 10 participants. Inter– and within– participant SD are also reported for susceptibility and volume in all hippocampal subregions.

| SEGMENT | SUSCEPTIBILITY (ppm) |  |  | VOLUME (mm<sup>3</sup>) |  |  |
| --- | --- | --- | --- | --- | --- | --- |
|  | Mean | Inter-participant SD | Within-participant SD across repeats | Mean | Inter-participant SD | Within-participant SD across repeats |
| ERC | 0.0047 | 0.0045 | 0.0019 | 748 | 116 | 71 |
| SUB | 0.0041 | 0.0023 | 0.0020 | 1204 | 153 | 40 |
| DG | -0.0133 | 0.0058 | 0.0023 | 1561 | 168 | 33 |
| TAIL | -0.0119 | 0.0082 | 0.0040 | 194 | 57 | 33 |
| CA1 | -0.0035 | 0.0034 | 0.0017 | 2940 | 438 | 80 |
| CA2 | -0.0026 | 0.0156 | 0.0084 | 15 | 7 | 3 |
| CA3 | -0.0092 | 0.0085 | 0.0055 | 53 | 18 | 6 |

*SD, Standard deviation; ERC, entorhinal cortex; SUB, subiculum; DG, dentate gyrus; TAIL, tail of the hippocampus; CA1, Cornu Ammonis 1; CA2, Cornu Ammonis 2; CA3, Cornu Ammonis 3.*

The pattern of susceptibility variation across subfields reported here is consistent with a previously published 7T study evaluating the effects of Alzheimer’s disease and post-SARS-CoV2 infection on hippocampal subfield susceptibility, which used ASHS for segmentation but used dual-echo GRE images with anisotropic voxel dimensions for QSM (26). It is also largely in agreement with a recent 7T study comparing QSM measurements in participants with MCI to healthy controls (25–27), which reported susceptibility values varying as SUB > CA1> CA3 > CA2 > DG spanning a range of approximately 20 ppb (the ERC and TAIL subfields were not evaluated). That study used a different approach for subfield segmentation (the deep learning-based Hippocampal Segmentation Factory tool) (38) and a significantly different 0.5 mm isotropic resolution three-echo GRE acquisition for QSM, but similar differences in susceptibility across the subfields were reported, suggesting that this is a general finding. A different referencing strategy was also used in the study by Zheng *et al*. (27) with susceptibility values referenced to the whole hippocampus susceptibility, but this yielded subfield susceptibility measurements similar to those reported in our study in which whole-brain referencing was used. We did also evaluate the effect of referencing to manually positioned ROIs within the ventricular cerebrospinal fluid (CSF). However, we found that this approach increased the variability of the susceptibility measurements (Supporting Information Figure S2), as has been previously reported for the analysis of similar data for a range of larger ROIs (5).

The relatively small values of the standard deviations across repeats, which are less than 4 ppb in the larger subfields and less than 8 ppb in the CA2 and CA3 subfields, give an indication of the level of susceptibility change that could be tracked longitudinally over time in individual subjects, while the larger inter-participant standard deviations provide guidance for estimating the magnitude of subfield susceptibility differences that can be measured between groups with a similar age range to the cohort studied here. For context, Zheng *et al.* found a statistically significant difference of susceptibility in the left DG subfield of 3 ppb between a group of 13 elderly participants with MCI (–16 ppb) versus 13 normal controls (–13 ppb) (27).

The subfield volumes reported in Figures 3 & 4 and in Table 1 are consistent with values expected from use of ASHS with the UMC Utrecht 7T ASHS Atlas (39), showing a large CA1 volume, and much smaller volumes for the CA2 and CA3 subfields. The average values of the standard deviations across repeats range from 80 mm^3^ in CA1 (2.7% of the average CA1 volume) to 3 mm^3^ in CA2 (20% of the average CA2 volume). In the larger subfields (ERC, SUB, DG, TAIL and CA1) the ratio of within participant standard deviation to average value is largest in the TAIL (17%). The inter-participant standard deviations are more than 1.6 times larger than the within-participant standard deviations for all subfields, and the ICC(2,k) values are correspondingly greater than 0.87 in all subfields, highlighting the capability to characterise small differences in subfield volumes via hippocampal segmentation of high-resolution, T1 and T2-weighted images acquired at 7T (22,35,40,41).

Limitations of the present study include the use of a single, rather than multi-echo, GRE acquisition for QSM. The data set that we analysed was acquired around 8 years ago and the choice of sequences to use in the UK7T travelling head study were consequently made prior to the consensus recommendation of the use of a monopolar multi-echo acquisition (42). It would be valuable in future to repeat the study using the approach set out in the consensus paper (42), and also to acquire data in older healthy participants or participants with MCI or early AD who might be expected to undergo a greater degree of movement during scanning. Movement would be expected to increase the variability of measured susceptibility values, while the use of a multi-echo sequence would reduce sensitivity to non-susceptibility related sources of phase variation and likely provide an increased signal-to-noise ratio in the field measurements. The agreement of the mean susceptibility values which we report with those described in studies using multi-echo acquisitions (26,27) provides some evidence that the sequence related differences do not significantly confound the findings reported here.

## Data Availability

All data produced in the present study are available upon reasonable request to the authors

## ACKNOWLEDGEMENTS

Associated Funding: MRC MR/N008537/1, MRC MR/T005580/1, UKRI/EPSRC EP/Y015398/1. OFA thanks the TETFund Nigeria and the University of Abuja for the invaluable support and opportunity to undertake this research as part of his PhD programme. We thank other members of the UK7T consortium (Ian Driver, Richard Wise, Tyler Morgan, Keith Muir, Will Clarke and Stuart Clare) for their contribution to the acquisition of data for the travelling head study.

## SUPPORTING INFORMATION

**Table S1:** Scanner systems used for data acquisition at each site involved in the UK7T travelling head study.

| **Subjects** | **Location** | **Vendor** | **Scanner Model** | **Software** |
| --- | --- | --- | --- | --- |
| 1&2 | Wellcome Centre for Integrative Neuroimaging (FMRIB), University of Oxford | Siemens | Magnetom 7T | VB17A |
| 3&4 | Cardiff University Brain Research Imaging Centre, Cardiff University | Siemens | Magnetom 7T | VB17A |
| 5&6 | Sir Peter Mansfield Imaging Centre, University of Nottingham | Philips | Achieva 7T | R5.1.7.0 |
| 7&8 | Wolfson Brain Imaging Centre, University of Cambridge | Siemens | Magnetom Terra | VE11U |
| 9&10 | Imaging Centre of Excellence, University of Glasgow | Siemens | Magnetom Terra | VE11U |

**Figure S1.**
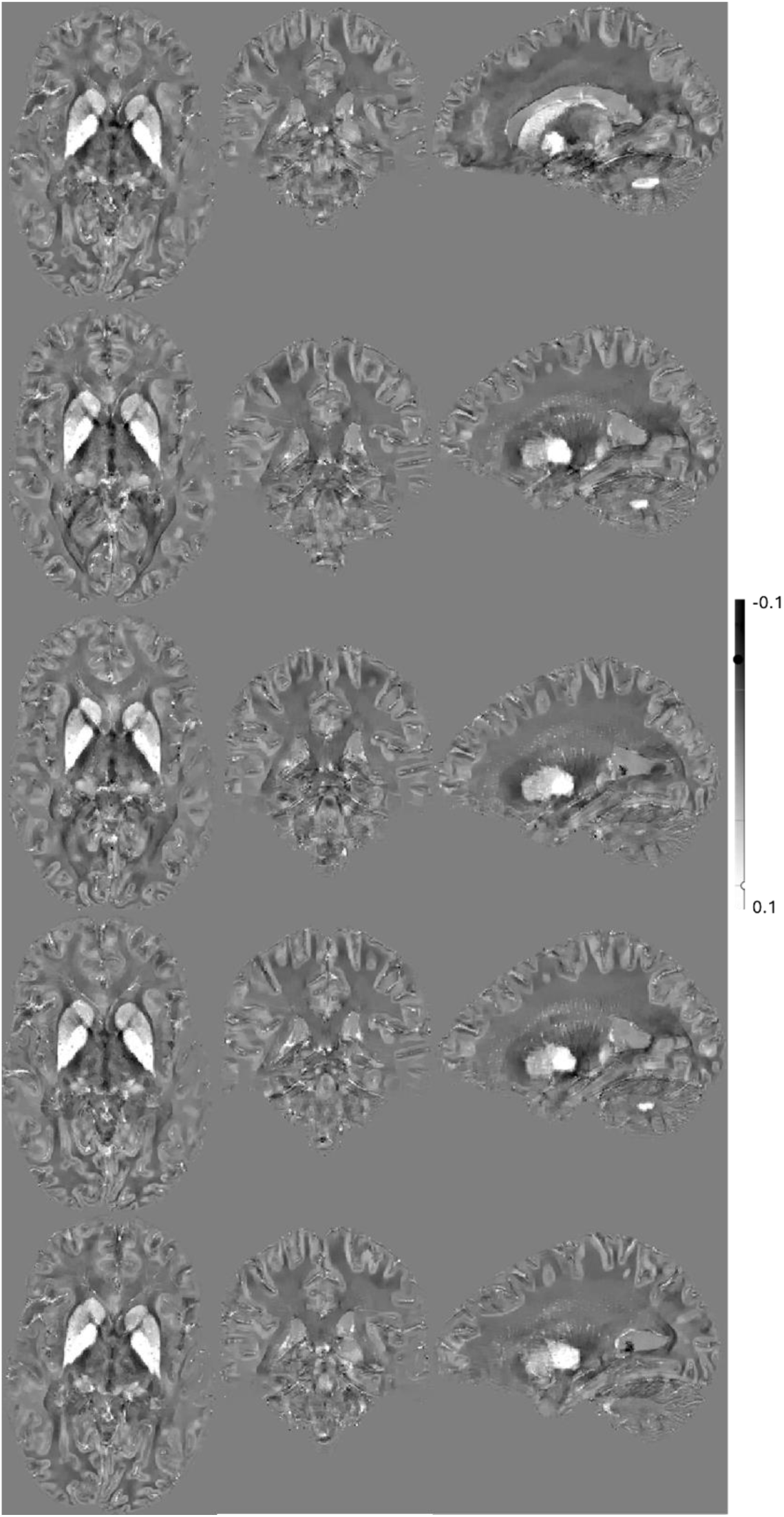
QSM data from a single participant acquired across five repeated scans.

**Figure S2.**
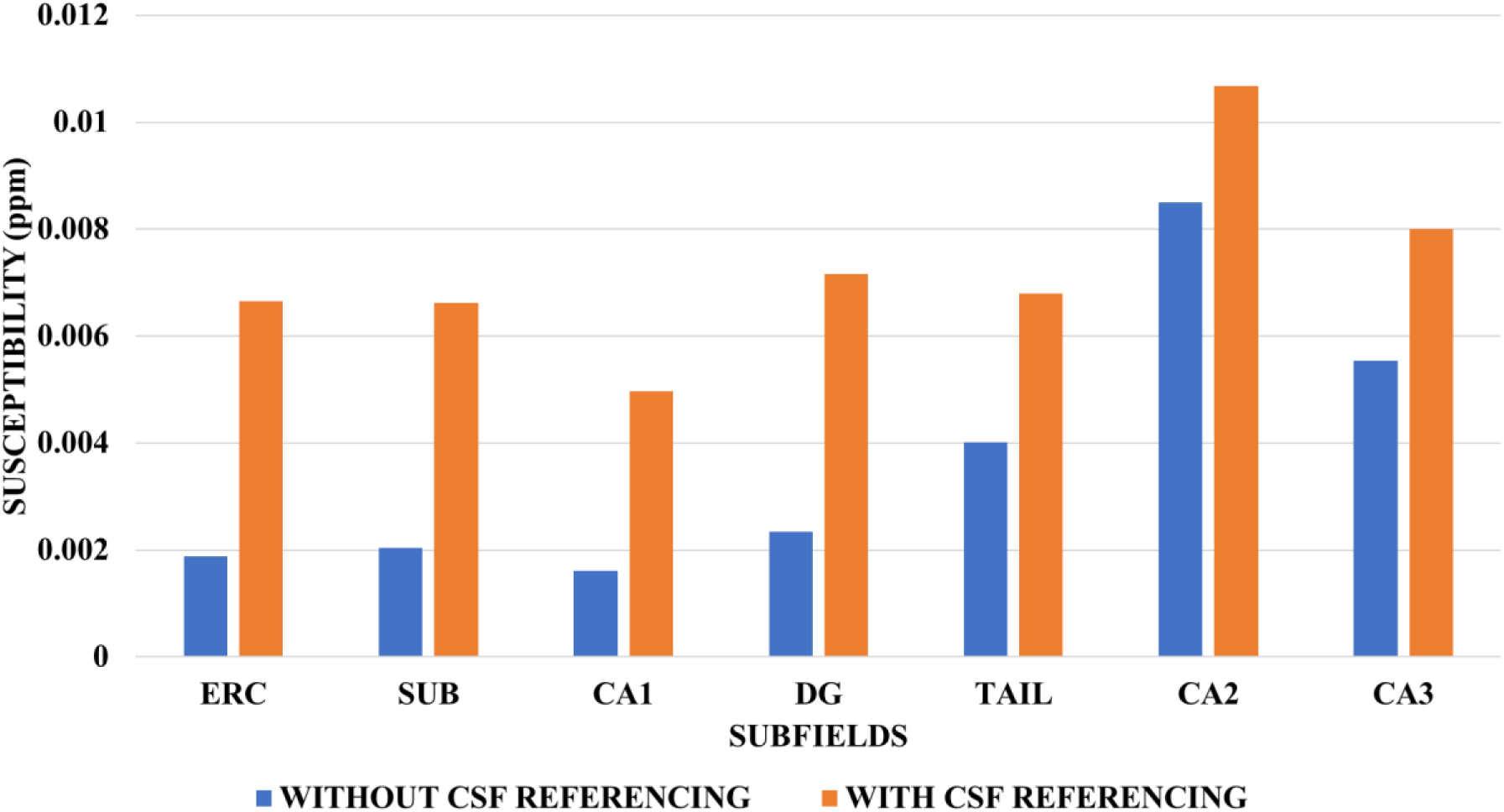
Mean within-participant standard deviation (SD) of susceptibility across five repeated measurements, averaged over the 10 participants, with (orange), and without (blue) CSF referencing.

**Table S3.** – Intraclass correlation coefficients (ICCs) for hippocampal subfield volume and susceptibility across repeated measurements.

| Subfield /Measure | ICC(2,1) Susceptibility | ICC(2,k) Susceptibility | ICC(2,1) Volume | ICC(2,k) Volume |
| --- | --- | --- | --- | --- |
| CA1 | 0.71 | 0.92 | 0.95 | 0.99 |
| CA2 | 0.68 | 0.92 | 0.72 | 0.93 |
| CA3 | 0.48 | 0.82 | 0.88 | 0.97 |
| DG | 0.83 | 0.96 | 0.96 | 0.99 |
| ERC | 0.84 | 0.96 | 0.57 | 0.87 |
| SUB | 0.48 | 0.82 | 0.91 | 0.98 |
| TAIL | 0.77 | 0.94 | 0.71 | 0.92 |

**Abbreviations:** ICC(2,1) and ICC(2,k) intraclass correlation coefficients for susceptibility and volume calculated using two-way random-effects models (single- and average-measure agreement, respectively).

**Table S4.** The statistical significance of the differences in susceptibility in ppm between the different subfields was tested using a one-way repeated ANOVA (setting the alpha value to 0.05), followed by Tukey’s Honest Significant Difference (HSD) test. Significant differences are indicated with an asterisk.

| SUBFIELD | SUBFIELD | Mean Difference | Std. Error | Sig |
| --- | --- | --- | --- | --- |
| ERC | SUB | 0.001 | 0.002 | 0.697 |
|  | DG | .018* | 0.003 | <.001 |
|  | TAIL | .017* | 0.003 | <.001 |
|  | CA1 | .008* | 0.001 | <.001 |
|  | CA2 | 0.007 | 0.005 | 0.174 |
|  | CA3 | .014* | 0.003 | 0.001 |
| SUB | ERC | -0.001 | 0.002 | 0.697 |
|  | DG | .017* | 0.002 | <.001 |
|  | TAIL | .016* | 0.002 | <.001 |
|  | CA1 | .008* | 0.001 | <.001 |
|  | CA2 | 0.007 | 0.004 | 0.129 |
|  | CA3 | .013* | 0.002 | <.001 |
| DG | ERC | -.018* | 0.003 | <.001 |
|  | SUB | -.017* | 0.002 | <.001 |
|  | TAIL | -0.001 | 0.002 | 0.58 |
|  | CA1 | -.010* | 0.002 | <.001 |
|  | CA2 | -.011* | 0.003 | 0.012 |
|  | CA3 | -.004* | 0.002 | 0.022 |
| TAIL | ERC | -.017* | 0.003 | <.001 |
|  | SUB | -.016* | 0.002 | <.001 |
|  | DG | 0.001 | 0.002 | 0.58 |
|  | CA1 | -.008* | 0.002 | 0.007 |
|  | CA2 | -.009* | 0.004 | 0.032 |
|  | CA3 | -0.003 | 0.002 | 0.246 |
| CA1 | ERC | -.008* | 0.001 | <.001 |
|  | SUB | -.008* | 0.001 | <.001 |
|  | DG | .010* | 0.002 | <.001 |
|  | TAIL | .008* | 0.002 | 0.007 |
|  | CA2 | -0.001 | 0.004 | 0.832 |
|  | CA3 | .006* | 0.002 | 0.019 |
| CA2 | ERC | -0.007 | 0.005 | 0.174 |
|  | SUB | -0.007 | 0.004 | 0.129 |
|  | DG | .011* | 0.003 | 0.012 |
|  | TAIL | .009* | 0.004 | 0.032 |
|  | CA1 | 0.001 | 0.004 | 0.832 |
|  | CA3 | .007* | 0.003 | 0.048 |
| CA3 | ERC | -.014* | 0.003 | 0.001 |
|  | SUB | -.013* | 0.002 | <.001 |
|  | DG | .004* | 0.002 | 0.022 |
|  | TAIL | 0.003 | 0.002 | 0.246 |
|  | CA1 | -.006* | 0.002 | 0.019 |
|  | CA2 | -.007* | 0.003 | 0.048 |

## REFERENCES

1. Wang Y, Liu T. Quantitative susceptibility mapping (QSM): Decoding MRI data for a tissue magnetic biomarker. Magn Reson Med. 2015;73(1):82–101.

2. Langkammer C, Schweser F, Krebs N, Deistung A, Goessler W, Scheurer E, et al. Quantitative susceptibility mapping (QSM) as a means to measure brain iron? A post mortem validation study. Neuroimage. 2012 Sep;62(3):1593–9.

3. Deistung A, Schäfer A, Schweser F, Biedermann U, Turner R, Reichenbach JR. Toward in vivo histology: A comparison of quantitative susceptibility mapping (QSM) with magnitude-, phase-, and R2*-imaging at ultra-high magnetic field strength. Neuroimage. 2013 Jan 15;65:299–314.

4. Betts MJ, Acosta-Cabronero J, Cardenas-Blanco A, Nestor PJ, Düzel E. High-resolution characterisation of the aging brain using simultaneous quantitative susceptibility mapping (QSM) and R2* measurements at 7 T. Neuroimage. 2016 Sep 1;138:43–63.

5. Rua C, Clarke WT, Driver ID, Mougin O, Morgan AT, Clare S, et al. NeuroImage Multi-centre, multi-vendor reproducibility of 7T QSM and R2 ∗ in the human brain: Results from the UK7T study. Neuroimage. 2020;223(April) 117358.

6. Ward RJ, Zucca FA, Duyn JH, Crichton RR, Zecca L. The role of iron in brain ageing and neurodegenerative disorders. The Lancet Neurology. 2014. 13,p. 1045–60.

7. Cornett CR, Ehmann WD, Wekstein DR, Markesbery WR. Trace elements in Alzheimer’s disease pituitary glands. Biol Trace Elem Res. 1998;62(1–2):107–14.

8. Bartzokis G, Sultzer D, Cummings J, Holt LE, Hance DB, Henderson VW, et al. In vivo evaluation of brain iron in Alzheimer disease using magnetic resonance imaging. Arch Gen Psychiatry. 2000 Jan 1;57(1):47–53.

9. Bartzokis G, Cummings JL, Markham CH, Marmarelis PZ, Treciokas LJ, Tishler TA, et al. MRI evaluation of brain iron in earlier– and later-onset Parkinson’s disease and normal subjects. Magn Reson Imaging. 1999;17(2):213–22.

10. Bunzeck N, Singh-Curry V, Eckart C, Weiskopf N, Perry RJ, Bain PG, et al. Motor phenotype and magnetic resonance measures of basal ganglia iron levels in Parkinson’s disease. Park Relat Disord. 2013 Dec;19(12):1136–42.

11. Sofic E, Paulus W, Jellinger K, Riederer P, Youdim MBH. Selective Increase of Iron in Substantia Nigra Zona Compacta of Parkinsonian Brains. J Neurochem. 1991;56(3):978–82.

12. Ravanfar P, Loi SM, Syeda WT, Van Rheenen TE, Bush AI, Desmond P, et al. Systematic Review: Quantitative Susceptibility Mapping (QSM) of Brain Iron Profile in Neurodegenerative Diseases. Front Neurosci. 2021; 18;15:618435

13. Uchida Y, Kan H, Sakurai K, Oishi K. Quantitative susceptibility mapping as an imaging biomarker for Alzheimer’s disease: The expectations and limitations. Front Neurosci. 2022 Aug 5;16:938092.

14. Chen L, Soldan A, Faria A, Albert M, Zijl PCM Van, Li X. Susceptibility MRI predicts mild cognitive impairment onset and cognitive decline in cognitively unimpaired older adults. Radiology. 2025 Sep;316(3):e250513.

15. Ayton S, Fazlollahi ÃA, Bourgeat ÃP, Raniga P, Ng A, Lim YY, et al. Cerebral quantitative susceptibility mapping predicts amyloid–b –related cognitive decline. Brain. 2017;140(8):2112–9.

16. Du L, Zhao Z, Cui A, Zhu Y, Zhang L, Liu J, et al. Increased Iron Deposition on Brain Quantitative Susceptibility Mapping Correlates with Decreased Cognitive Function in Alzheimer’s Disease. ACS Chem Neurosci. 2018 Jul 18;9(7):1849–57.

17. Manjón J V., Romero JE, Coupe P. A novel deep learning based hippocampus subfield segmentation method. Sci Rep. 2022;12(1):1–9.

18. Bartsch T. The Clinical Neurobiology of the Hippocampus: An Integrative View OUP Oxford; 2012.

19. Braak H, Braak E. Neuropathological stageing of Alzheimer-related changes. Acta Neuropathol. 1991;82(4):239–59.

20. Jack R, Bentley D, Twomey K, Zinsmeister R. MR imaging-based volume measurements of the hippocampal formation and anterior temporal lobe: validation studies. Radiology. 1990 Jul;176(1):205–9

21. Schuff N, Woerner N, Boreta L, Kornfield T, Shaw LM, Trojanowski JQ, et al. MRI of hippocampal volume loss in early Alzheimers disease in relation to ApoE genotype and biomarkers. Brain. 2009;132(4):1067–77.

22. Adeyemi OF, Gowland P, Bowtell R, Mougin O, Hosseini AA. Hippocampal Subfield Volume in Relation to Cerebrospinal Fluid Amyloid-ß in Early Alzheimer’s Disease: Diagnostic Utility of 7T MRI. Eur J Neurol. 2025;32(2):1–11.

23. Kim HG, Park S, Rhee HY, Lee KM, Ryu CW, Rhee SJ, et al. Quantitative susceptibility mapping to evaluate the early stage of Alzheimer’s disease. NeuroImage Clin. 2017;16:429–38.

24. Kan H, Yuto U, Arai N, Ueki Y, Aoki T, Kasai H, et al. Simultaneous voxel-based magnetic susceptibility and morphometry analysis using magnetization-prepared spoiled turbo multiple gradient echo. NMR Biomed. 2020 May;33(5):e4272.

25. Kagerer SM, Vionnet L, Bergen JMG Van, Meyer R, Gietl AF, Pruessmann KP, et al. Hippocampal iron patterns in aging and mild cognitive impairment. Front Aging Neurosci. 2025 Jul 2;17:1598859.

26. Hosseini A, Adeyemi O, Bowtell R, Gowland P, Ibrahim T, Liou JJ, et al. Using 7T MRI to study hippocampal structures in Alzheimer’s disease and post-SARS-CoV2 infection. Alzheimer’s Dement. 2023 Dec 25;19.

27. Zheng G, Yu FF, Huang Y, Mazumder S, Vargas SL, Unschuld PG, et al. Hippocampal Subfield Susceptibility Alterations in Mild Cognitive Impairment Revealed by 7T MRI. AJNR Am J Neuroradiol. 2026 May 4;47(5):1308–1314.

28. Deh K, Nguyen TD, Eskreis-Winkler S, Prince MR, Spincemaille P, Gauthier S, et al. Reproducibility of quantitative susceptibility mapping in the brain at two field strengths from two vendors. J Magn Reson Imaging. 2015 Dec 1;42(6):1592–600.

29. Naji N, Lauzon ML, Frayne R, Lebel C, Seres P, Stolz E, et al. Multisite reproducibility of quantitative susceptibility mapping and effective transverse relaxation rate in deep gray matter at 3 T using locally optimized sequences in 24 traveling heads. NMR Biomed. 2022 Nov;35(11):e4788.

30. Clarke WT, Mougin O, Driver ID, Rua C, Morgan AT, Asghar M, et al. Multi-site harmonization of 7 tesla MRI neuroimaging protocols. Neuroimage. 2020 Feb 1;206:116335.

31. Acosta-Cabronero J, Milovic C, Mattern H, Tejos C, Speck O, Callaghan MF. A robust multi-scale approach to quantitative susceptibility mapping. Neuroimage. 2018 Dec 1;183:7–24.

32. Zhou D, Liu T, Spincemaille P, Wang Y. Background field removal by solving the Laplacian boundary value problem. NMR Biomed. 2014 Mar;27(3):312–9.

33. Li W, Wu B, Liu C. Quantitative susceptibility mapping of human brain reflects spatial variation in tissue composition. Neuroimage. 2011 Apr 15;55(4):1645–56.

34. Yushkevich PA, Pluta JB, Wang H, Xie L, Ding SL, Gertje EC, et al. Automated volumetry and regional thickness analysis of hippocampal subfields and medial temporal cortical structures in mild cognitive impairment. Hum Brain Mapp 2015 Jan 1;36(1):258–87.

35. Wisse LEM, Biessels GJ, Heringa SM, Kuijf HJ, Koek DL, Luijten PR, et al. Hippocampal subfield volumes at 7T in early Alzheimer’s disease and normal aging. Neurobiol Aging. 2014;35(9):2039–45.

36. Jenkinson M, Beckmann CF, Behrens TEJ, Woolrich MW, Smith SM. FSL. NeuroImage Clin. 2011;62(2012):782–90.

37. Jenkinson M, Bannister P, Brady M, Smith S. Improved Optimization for the Robust and Accurate Linear Registration and Motion Correction of Brain Images. Neuroimage. 2002;17(2):825–41.

38. Poiret C, Bouyeure A, Patil S, Bottlaender M, Lemaitre F. A fast and robust hippocampal subfields segmentation: HSF revealing lifespan volumetric dynamics. Front Neuroinform. 2023 Jun 15;17:1130845.

39. Wisse LEM, Kuijf HJ, Honingh AM, Wang H, Pluta JB, Das SR, et al. Automated hippocampal subfield segmentation at 7T MRI. Am J Neuroradiol. 2016;37(6):1050–7.

40. Liou J jiun, Santini T, Li J, Gireud-goss M, Kautz TF, Parker-garza J, et al. BRAIN COMMUNICATIONS Examining neuroimaging biomarkers, plasma biomarkers and cognitive functions in patients with recovered COVID-19 infection: a multicentre study using 7T MRI. Brain Commun. 2026;8(2):1–13.

41. Kiran T, Marshall-gradisnik S, Eaton-fitch N, Markus B, Maira I, Barnden L. Hippocampal subfield volume alterations and associations with severity measures in long COVID and ME / CFS: A 7T MRI study. PLoS One.2025 Jan 13;20(1):e0316625.

42. Bilgic B, Lee J, Li X, Liu C, Marques JP, Milovic C. Recommended Implementation of Quantitative Susceptibility Mapping for Clinical Research in The Brain: A Consensus of the ISMRM Electro-Magnetic Tissue Properties Study Group. Magn Reson Med. 2025;91(5):1834–62.

